# Analyzing changes in respiratory rate to predict the risk of COVID-19 infection

**DOI:** 10.1101/2020.06.18.20131417

**Authors:** Dean J. Miller, John V. Capodilupo, Michele Lastella, Charli Sargent, Gregory D. Roach, Victoria H. Lee, Emily R. Capodilupo

## Abstract

COVID-19, the disease caused by the SARS-CoV-2 virus, can cause shortness of breath, lung damage, and impaired respiratory function. Containing the virus has proven difficult, in large part due to its high transmissibility during the pre-symptomatic incubation. The study’s aim was to determine if changes in respiratory rate could serve as a leading indicator of SARS-CoV-2 infections. A total of 271 individuals (age = 37.3 ± 9.5, 190 male, 81 female) who experienced symptoms consistent with COVID-19 were included – 81 tested positive for SARS-CoV-2 and 190 tested negative; these 271 individuals collectively contributed 2672 samples (days) of data (1856 healthy days, 231 while infected with COVID-19 and 585 while infected with something other than COVID-19). To train a novel algorithm, individuals were segmented as follows; (1) a training dataset of individuals who tested positive for COVID-19 (n=57 people, 537 samples); (2) a validation dataset of individuals who tested positive for COVID-19 (n=24 people, 320 samples) ; (3) a validation dataset of individuals who tested negative for COVID-19 (n=190 people, 1815 samples). All data was extracted from the WHOOP system, which uses data from a wrist-worn strap to produce validated estimates of respiratory rate and other physiological measures. Using the training dataset, a model was developed to estimate the probability of SARS-CoV-2 infection based on changes in respiratory rate during night-time sleep. The model’s ability to identify COVID-positive individuals not used in training and robustness against COVID-negative individuals with similar symptoms were examined for a critical six-day period spanning the onset of symptoms. The model identified 20% of COVID-19 positive individuals in the validation dataset in the two days prior to symptom onset, and 80% of COVID-19 positive cases by the third day of symptoms.

## Introduction

The novel coronavirus disease (COVID-19) is caused by the severe acute respiratory syndrome coronavirus 2 (SARS-CoV-2) virus [1] and predominantly presents as a lower respiratory tract infection. Severe cases of the disease can result in alveolar damage and progressive respiratory failure [2]. Containing the virus has proven difficult due to its high transmissibility during the pre-symptomatic incubation phase [3], and widespread shortages of testing.

Outside of traditional laboratory testing, few practical COVID-19 monitoring systems have been proposed. Some businesses looking to reopen following physical distancing mandates have implemented daily monitoring of temperature to identify and isolate potentially infected individuals; while this method would be expected to have a high level of sensitivity – workers with fevers would correctly be sent home – infectious individuals that do not present with a fever may be exposed to colleagues during the 2 to 14-day pre-symptomatic incubation period [4, 5]. This limitation of fever-based screening is significant given that infectiousness is known to peak 0-2 days prior to symptom onset [6].

Respiratory rate is a common screening tool to identify lower respiratory tract infections in clinical settings [7]; guidelines define tachypnea as a respiratory rate greater than 20 respirations per minute (rpm), and advise further tests (e.g., chest radiography) when present [7]. While such thresholds are useful in clinical settings, they are only implemented once symptoms have emerged and are not sensitive to intraindividual differences in normal respiratory function. Given that COVID-19 impairs and damages the respiratory system [2], it is reasonable to suggest that changes in respiratory efficiency – and therefore resting respiratory rate – might occur in the early stages of infection. In this context, noninvasive daily monitoring of respiratory rate may be used to detect subclinical intraindividual deviations and identify potential infections that would otherwise be overlooked by clinical thresholds [7].

If deviations in respiratory rate are found to be an accurate indicator of COVID-19 infection, respiratory rate monitoring could form part of the protocol used by medical professionals and organizations to enforce self-isolation and target testing. The aim of this study was to assess the ability of a novel algorithm to classify changes in respiratory rate as indicative of COVID-19 infection immediately prior to and during the first days of symptoms and to evaluate the model’s robustness to instances of similar clinical presentations with differing etiology.

## Materials and methods

Respiratory rate, resting heart rate (RHR) and heart rate variability (HRV) were measured using the WHOOP strap; the algorithms used to derive these metrics from the wearable’s photoplethysmography sensor are beyond the scope of this paper, but have been validated in third party analysis and shown to have high levels of agreement with gold standard methodology [8]. The WHOOP strap is a small, waterproof, and rechargeable device containing a photoplethysmogram, accelerometer, thermometer, capacitive touch sensor, and gyroscope, which can be worn comfortably 24-hours per day and lasts 5 days between charges. The wrist-worn strap wirelessly transfers data to mobile devices running the associated WHOOP app; from there, data is transferred to a secure cloud-based data storage and processing server, collectively known as the *WHOOP system*.

The following physiological data were obtained from the WHOOP system for this study:

- Respiratory rate – median value of respirations per minute, derived each night during the main sleep period via photoplethysmography.
- RHR – average beats per minute sampled during the last five minutes of the last episode of slow wave sleep each night.
- HRV – sampled during the last five minutes of the last episode of slow wave sleep each night using the root mean square of successive RR interval differences (rMSSD) method in units of milliseconds.

In addition to automated tracking of physiological data, the WHOOP app supports tracking of manually reported contextual factors. In response to the COVID-19 pandemic, on 14 March 2020, WHOOP added the ability to track COVID-19 symptoms and test results. Member-reported incidences of COVID-19 symptoms and test results were extracted through 06 June 2020. Respiratory rate, RHR, and HRV were extracted between 01 November 2019 and 30 November 2019; respiratory rate was additionally sampled between 1 January 2020 and 06 June 2020 for individuals that reported test results for COVID-19.

## Data analysis

### Stability of metrics

Respiratory rate was evaluated as a potentially sensitive indicator of infection due to anecdotal observations of low internight variation in WHOOP data. A search of Pubmed showed no extant longitudinal studies reporting on variability of nightly respiratory rate in healthy adults. Therefore, to support the use of this metric in the model, a supplementary dataset from November 2019 was generated for analysis. A date range of 1 November 2019 through 30 November 2019 was chosen to avoid confounding factors related to the COVID-19 pandemic. A total of 25,000 WHOOP members were randomly selected (n = 750,000 nights); the only inclusion criteria was having respiratory rate recorded on all 30 consecutive nights. Resting heart rate and resting heart rate variability over this period were included for comparison. The following variables were calculated from the November dataset for each of the physiological metrics:

- Mean intraindividual mean: *mean within-member means*.
- Standard deviation of intraindividual means: *standard deviation of within-member means*.
- Mean intraindividual standard deviation: *mean within-member standard deviation*.
- Standard deviation of intraindividual standard deviations: *standard deviation of within-member standard deviations*.
- Coefficient of variation: *intraindividual standard deviation divided by the intraindividual mean*.

### Predictive model

#### Data Extraction

A total of 271 adults (age = 37.3 ± 9.5, 190 male, 81 female) were included in the study; inclusion criteria were (1) self-reporting symptoms consistent with COVID-19 (i.e., cough, fever and/or fatigue) and (2) having been tested for the SARS-CoV-2 virus. These individuals were separated into three groups:

- training dataset: *individuals who began experiencing COVID-19 symptoms between 14 March 2020 and 14 April 2020 (n = 57);*
- validation dataset 1: *COVID-19 positive individuals who began experiencing COVID-19 symptoms between 14 April and 6 June 2020 (n = 24);*
- validation dataset 2: *individuals who experienced COVID-19 symptoms but reported a negative test result (n = 190)*.

In order to develop the algorithm, data were categorized by day relative to symptom onset (day 0) into:

- healthy days: *data extracted from 30 to 14 days prior to symptom onset;*
- infected days: *data extracted between 2 days prior to symptom onset and 3 days post symptom onset*.

All 271 individuals contributed to both categories, with a maximum of 15 healthy days per person and 6 infected days per person. For the training dataset, 146 infected days and 391 healthy days were included. Due to the class imbalance between infected days and healthy days, synthetic samples (i.e., days) were generated for the positive class (i.e., infected days) by adding uniformly distributed random noise on the interval [0, 1) to each infected day, bringing the number of infected days to 292. Generation of synthetic samples was done only for training and was not repeated for the validation datasets. Synthetic samples were only used for training the model and were excluded from the analysis of the training set presented throughout. For validation dataset 1, 85 infected days and 235 healthy days were included. For validation dataset 2, 585 infected days and 1230 healthy days were included.

#### Data transformation

The daily respiratory rate value (herein, *current value*) for each individual was transformed into features based on how it compared to the values taken on each of the 21 days prior. In all datasets, only current values for which the prior 21 consecutive nights’ respiratory rates were available were included. These features capture the dynamics of deviation from recent trends along a variety of time scales. In generating the classifier’s features, the following metrics were used:

- **RR**_**0**_: *current value (a respiratory rate)*
- 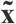: *median of the respiratory rates in the 14 day period between 21 and 7 nights prior to the current value*.
- **σ**: *standard deviation of the respiratory rates in the 14 day period between 21 and 7 nights prior to the current value*.
- **μ**_**2**_: *mean of the current value and immediately prior night’s respiratory rate*.
- **μ**_**3**_: *mean of the current value and immediately prior two nights’ respiratory rates*.
- **μ**_**6**_: *median of the immediately prior 6 nights of respiratory rates, excluding* **RR**_**0**_.
- **m**_**6**_: *slope of the linear regression of the collection of the respiratory rates of the current day to 6 days prior, excluding* **RR**_**0**_.

The features derived from these metrics were:

1. 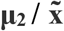
2. 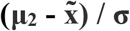
3. **RR**_**0**_ **-μ**_**3**_
4. **m**_**6**_
5. **μ**_**2**_ **-μ**_**6**_

Collectively, these internally derived and novel features capture dynamics of the changes in respiratory rate over time. Utilizing a modified z-score (i.e., utilizing a median value rather than mean), creates a baseline that is robust to outlier values and more stable over the short time periods explored in this study. Using a lagged baseline, as in 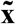 in features 1 and 2, allows data to increase during an incubation period without artificially elevating the baseline and masking the impact of the SARS-CoV-2 infection.

A gradient boosted classifier was trained using Python Language Software (version 3.6.2) on the derived features to return a probability of SARS-CoV-2 infection on healthy and infected days.

#### Model performance

In order to evaluate the model’s performance for classifying healthy and infected days, a threshold value was assigned to the probability output of the model such that meeting or exceeding that threshold was equivalent to classifying healthy or infected days as COVID-19 positive (C+); while failing to exceed the threshold was equivalent to classifying healthy or infected days as COVID-19 negative (C-). The threshold value was strategically set at 0.3 to maximize the utility of the model by reducing the chance of false negatives at the expense of increasing false positives, in recognition that false negatives may have higher costs to society than false positives. The model’s performance for classifying healthy days and infected days for each dataset was also evaluated at that threshold by calculating sensitivity, specificity, positive predictive value (PPV) and negative predictive value (NPV).

## RESULTS

### Stability of metrics

Thirty-day intraindividual variability of metrics are presented in **Table 1**. Respiratory rate was found to have a lower coefficient of variation than both heart rate variability and resting heart rate.

**Table 1.** Intraindividual Means and Standard Deviations for selected metrics.

| Metric | Intraindividual Mean (M±SD) | Intraindividual SD (M±SD) | Coefficient of Variation (%) |
| --- | --- | --- | --- |
| Respiratory rate (rpm) | 15.53 ± 1.42 | 0.51 ± 0.20 | 3.28% |
| Resting heart rate (bpm) | 55.89 ± 7.37 | 4.83 ± 1.77 | 8.60% |
| Heart rate variability (ms) | 65.22 ± 30.86 | 17.74 ± 9.59 | 27.20% |

### Predictive model

The model returned a continuous probability that a given sample is indicative of a SARS-CoV-2 infection (**Fig 1). Table 2** summarizes the performance of this model after mapping the model’s continuous probability output into C+ and C-classifications, bifurcated on the threshold of 0.3.

**Table 2.** Performance of the model for the classification of healthy days and infected days for each dataset.

| **Dataset** | **Sensitivity** | **Specificity** | **PPV** | **NPV** |
| --- | --- | --- | --- | --- |
| Training dataset - COVID-19 Positive | 41.1% | 98.5% | 90.9% | 81.7% |
| Validation dataset 1 - COVID-19 Positive | 36.5% | 95.3% | 73.8% | 80.6% |
| Validation dataset 2 - COVID-19 Negative | 17.1% | 95.0% | 61.7% | 70.7% |

Note: This table evaluates the model's ability to discriminate between healthy days and infected days for each dataset. In the training and first validation dataset, all infected days are COVID-19 positive while the second validation dataset's infected days are COVID-19 negative.

**Fig 1.**
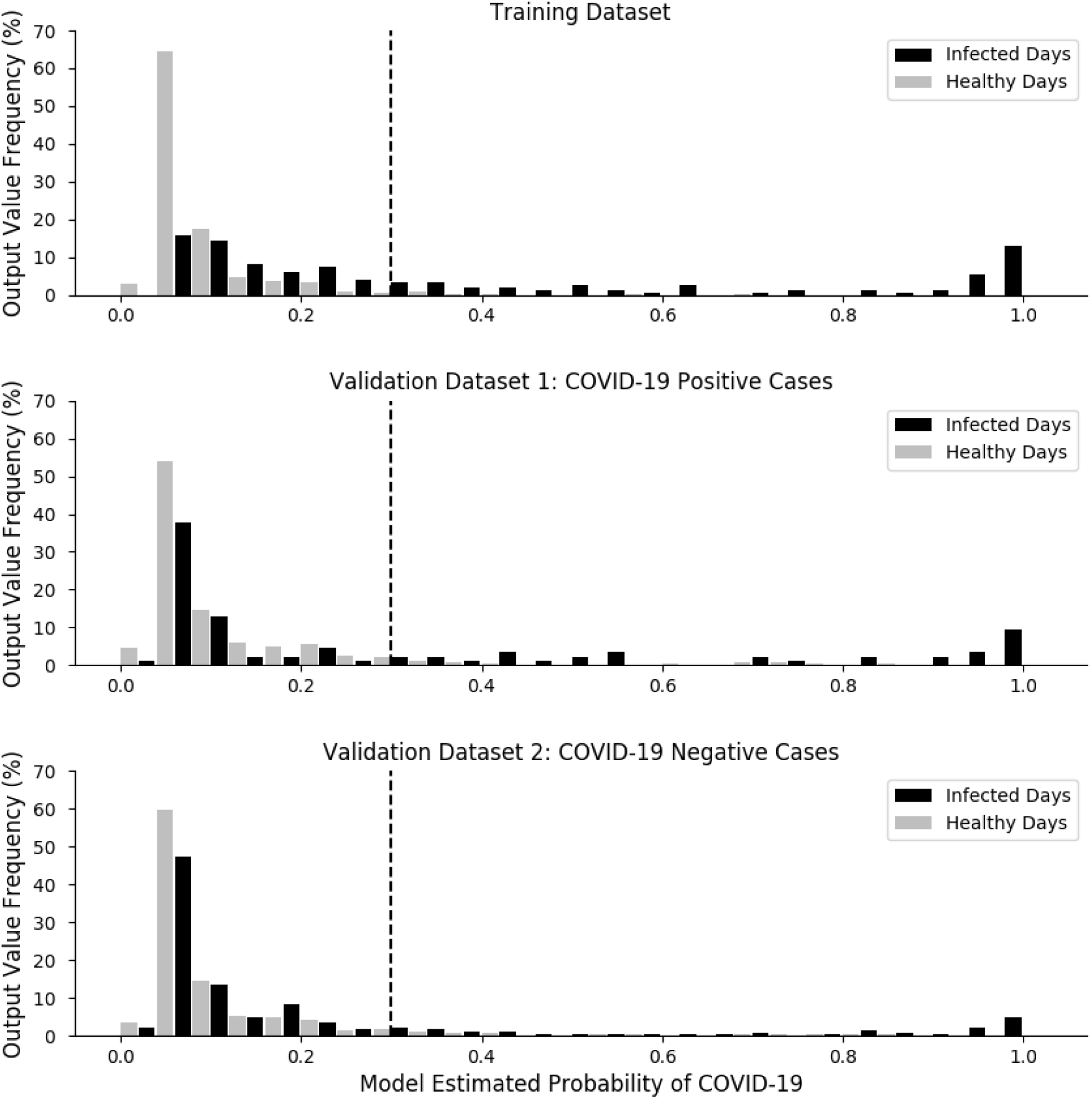
Top panel: Distribution of classifier output from the training dataset, separating infected days and healthy days. Middle panel: Distribution of the classifier output from the validation dataset separating COVID-19 positive infected days and healthy days. Bottom panel: Distribution of the classifier output from the validation dataset separating COVID-19 negative infected days and healthy days. For all panels, black bars denote infected days (top and middle: COVID-19 positive, bottom: COVID-19 negative), gray bars denote healthy days; y-axes show the relative frequency of the classifier’s output values, binned with widths of 0.04, with infected days and healthy bars plotted side by side. Dashed black lines show the threshold value of 0.3 which is used to map the continuous distribution of the model’s output to a binary classification.

**Fig 2** quantifies the percentage of individuals in each dataset to whom the model would have assigned a positive or negative COVID-19 classification relative to symptom onset. Note that only the subset of users who had available samples on each of the 6 evaluated days were included. In the training dataset (n=23), 73.9% of individuals had at least one correct C+ classification over the six day period. In validation dataset 1 (n=10), 80.0% of the individual subjects had at least one correct C+ classification; during that same relative time period, while 34.2% of the individuals in validation dataset 2 (n=79) had one or more C+ classification.

**Fig 2.**
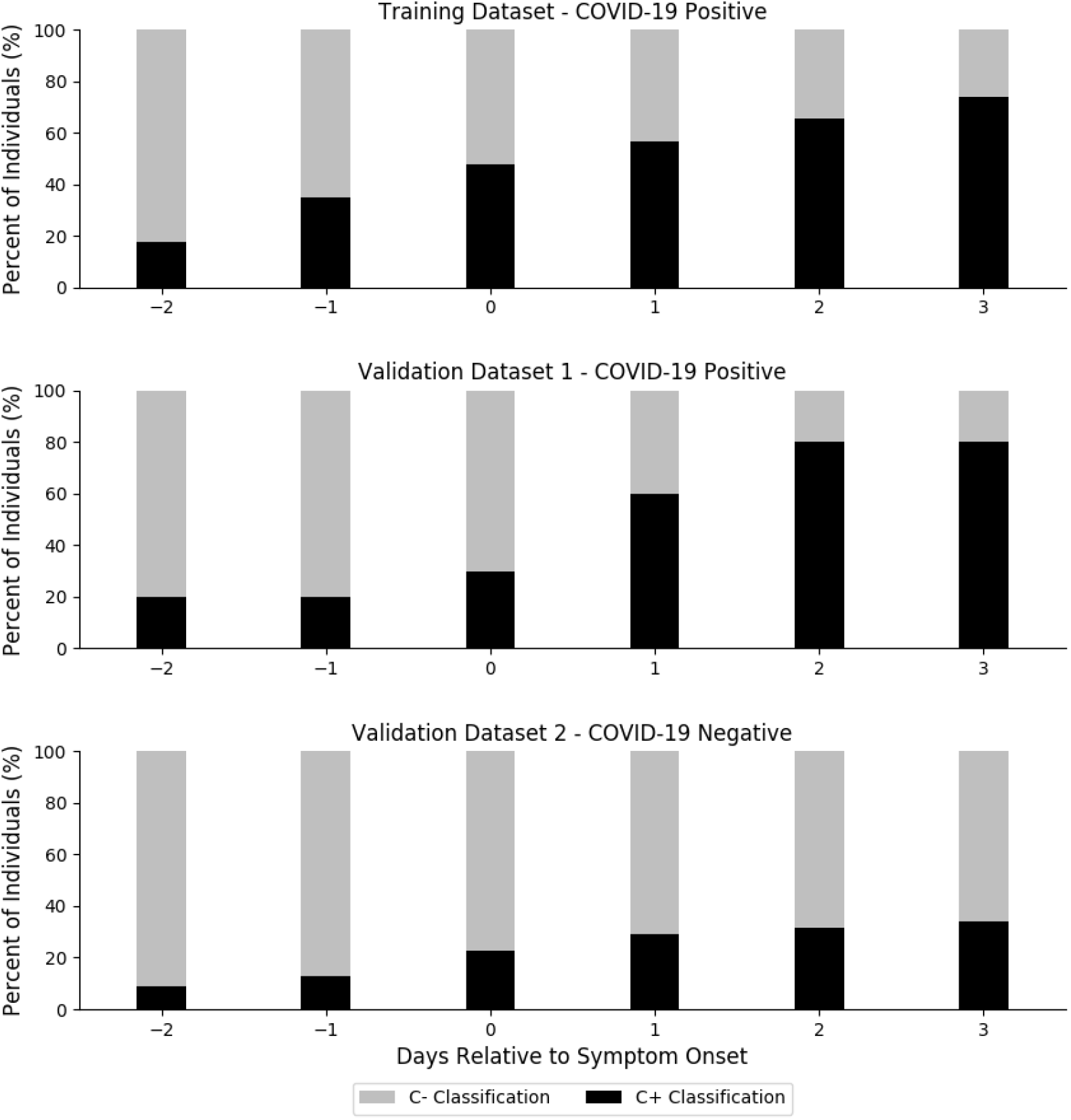
Cumulative percentage of individuals from each dataset that were classified as COVID-19 positive (C+) or COVID-19 negative (C-) relative to symptom onset (day=0).

## DISCUSSION

The aim of this study was to assess the ability of a novel algorithm to classify changes in respiratory rate, as indicative of COVID-19 infection immediately prior to and during the first days of symptoms. The major findings of this study are (1) stability of nightly respiratory rate measurements within healthy individuals makes it a useful metric for tracking changes in wellness; (2) the model is capable of distinguishing between healthy days and infected days for individuals that tested positive to COVID-19 as well as those who had symptoms but tested negative; (3) the model identified 20% of individuals of COVID-19 positive prior to the onset of symptoms, and correctly identified 80% of COVID-19 positive individuals by the third day of symptoms.

### Stability of metrics

This is the first study to report on nightly changes in resting heart rate, heart rate variability, and respiratory rate in healthy individuals. Our findings show that while interindividual variation in nightly respiratory rate can be large, intraindividual variability across 30 nights is typically quite small, with mean intraindividual standard deviation of 0.51 ± 0.20 rpm. The finding that nighttime median respiratory rate in healthy individuals has low internight variability is a novel finding of this paper and suggests that deviations in respiratory rate may be a useful indicator of acute changes in lower respiratory tract health.

### Predictive model

This is the first study to examine the potential for continuously monitored respiratory rate to identify early stages of COVID-19 infections. A predictive algorithm was formulated to leverage individual baseline data and determine if nightly respiratory rate when contextualized by 21-day trends can predict COVID-19 infections. A significant finding was that 20% of COVID-19 positive individuals were identified prior to the onset of symptoms and 80% of COVID-19 positive individuals were correctly identified by the third day of symptoms (**Fig 2**). This suggests that the final stages of incubation and early stages of the infection may have a detectable signature that can identify individuals who should self-isolate and seek testing. This novel approach may be particularly advantageous for individuals with low resting respiratory rates, who despite experiencing significantly elevated respiratory rates relative to their personal baseline, might not be medically classified as tachypneic according to globally defined norms [7].

There are a number of practical applications for the current model’s ability to analyze daily changes in respiratory rate, including aiding testing protocols and monitoring essential workers. The limited availability of testing kits and the time-intensive nature of most laboratory tests makes repeated screening for an individual both costly and impractical. Despite strict testing criteria, 3-12% of laboratory COVID-19 tests return a positive result [9]. Given the performance of the current model at discriminating between COVID-19 and other illnesses with similar symptomatology, it could potentially be used to streamline testing protocols in areas that may have testing kit shortages. In addition, this algorithm may be particularly useful in situations where physical distancing is impractical (e.g., industry, elite sport, healthcare), but where a positive COVID-19 case could have major implications. Along with recommended hygiene and physical distancing protocols, wearable technology could be used as a point of care measure to monitor employees and/or athletes during the transition back to work and competition.

Some boundary conditions should be considered when interpreting the results of this study. Firstly, COVID-19 test results and date of symptom onset were reported by WHOOP members directly in the WHOOP app and were not verified by medical professionals. All COVID positive individuals included in the analyses experienced symptomatic COVID-19 disease, thus the model has not been evaluated for its performance in fully asymptomatic cases; given that asymptomatic COVID-19 cases are contagious [10], further analysis is required to determine the utility of the algorithm in those cases. We note that we did not collect final diagnoses from individuals who tested negative to COVID-19, so the COVID-19 negative cohort may represent individuals with a variety of illnesses; further research beyond the scope of this study is warranted to segment model performance by non-COVID-19 diagnosis, especially for conditions with similar initial clinical presentations. The number of unique individuals included in the analyses could be seen as a limitation, however the model was trained using data extracted from multiple days from each user.

When interpreting the model’s performance, it should be noted that the sensitivity and specificity of the model are determined both by the discriminatory power of the features and by the threshold selected to discriminate between C+ and C-designations. As illustrated in **Fig 1**, healthy days tend to be assigned lower probabilities of being COVID-19 positive while infected days tend to be assigned higher probabilities. For the same probability distributions, a higher threshold would result in higher specificity but lower sensitivity, while a lower threshold makes the opposite tradeoff increasing sensitivity while decreasing specificity. The optimal threshold for a given model is dependent on its intended application; while a threshold of 0.5 would maximize accuracy, this is often not the metric most associated with practical utility. For the algorithm presented in this study, a false positive – indicating that COVID-19 negative individual may be COVID-19 positive – means that an individual self-isolates unnecessarily, while a false negative – indicating that a COVID-19 positive individual is COVID-19 negative – could result in the individual interacting with and potentially infecting others. Therefore, the reduced threshold value of 0.3 was chosen in recognition that false negatives have higher costs to society than false positives. We note that the threshold selection process was not particularly rigorous and that the optimal tradeoff between false positives and false negatives would be dependent on a number of unknown factors beyond the scope of this analysis. Finally, it should be noted that the WHOOP strap is not a medical device and should not be used as a substitute for professional medical advice, diagnosis or treatment.

## Conclusions

This study presents a novel, non-invasive method for detecting SARS-CoV-2 infection prior to and during the first days of symptoms. The findings indicate that the early stages of the infection may have a detectable signature that could help identify individuals who should self-isolate and seek testing. Future investigations should examine the performance of respiratory rate based algorithms to classify infection among larger and more diverse cohorts.

## Data Availability

Data cannot be shared publicly because of user privacy.

## Acknowledgements

We thank Dr. Douglas Johnston and Dr. Aaron Weiss from the Cleveland Clinic for their help with this project.

